# Development and Evaluation of Heart Failure Patients Oral Comfort Assessment Questionnaire

**DOI:** 10.1101/2025.02.04.25321648

**Authors:** Mei Lu, Runyu Yang, Xiaona Li, Xinkun Wang, Ming Tao

## Abstract

**Background:** Our objective was to develop a reliable and valid questionnaire to assess the oral comfort status of inpatients with heart failure, providing medical staff with a scientifically grounded tool for evaluating their oral comfort.

**Methods:** A comprehensive pool of questionnaire items was developed through a literature review, qualitative interviews, and expert consultation. Six experts participated in two rounds of consultation to refine the items, followed by a pre-investigation involving 20 patients. Finally, 192 patients treated at Zun Yi Medical University between June and September 2020 were selected for project analysis and to assess the reliability and validity of the questionnaire.

**Results:** Through exploratory factor analysis, four common factors were identified, accounting for a cumulative variance contribution rate of 59.611%. The finalized questionnaire comprised 24 items, demonstrating robust reliability and validity. The Cronbach’s α coefficient was 0.810, indicating high internal consistency, while the Spearman-Brown coefficient was 0.732. The Pearson correlation coefficient was 0.233 (P < 0.05) when compared with the Beck Oral Scoring Method (BOAS table). The content validity index (CVI) for individual items ranged from 0.833 to 1. The overall content validity index (S-CVI/UA) for the questionnaire was 0.875, with an average content validity index (S-CVI/Ave) of 0.979.

**Conclusion:** The oral comfort assessment questionnaire developed in this study for inpatients with heart failure exhibits excellent reliability and validity, establishing it as an effective and valuable tool for assessing the oral comfort status of this patient population.

## Introduction

Heart failure (HF) is a severe or advanced stage of a variety of heart diseases with high morbidity and mortality. In the management of chronic diseases of heart failure, oral health has gradually received attention in recent years. A growing body of research ^[1-3]^ has emphasized the inclusion of oral health in heart failure care. “The Healthy Oral Action Plan 2019-2025^[4]^” issued by the National Health Commission clearly states that the oral health management of patients should be strengthened and the development of clinical oral health care should be encouraged. However, the current oral condition of hospitalized HF patients is not optimistic.

A variety of factors such as treatments and disease progression had a negative impact on the overall oral health of HF patients, including, the use of diuretics and restriction of fluid intake can lead to a decrease in saliva secretion ^[5]^. So, reducing the ability of the oral cavity to self-clean debris and bacteria, the patient’s self-care ability and insufficient activity tolerance has resulted in a decline in oral hygiene practices, leading to poor cleaning effectiveness. The bacterial proliferation leads to the acidic oral microenvironment of HF patients, and the activity of lysozyme in saliva in the acidic environment is weakened ^[6]^, which further reduces the bactericidal effect, promotes the reproduction of pathogenic bacteria, and leads to the dysbiosis of oral flora. These factors affect the oral comfort of HF patients, and patients with HF often report oral discomfort such as thick tongue coating, thirst, and taste changes, which in turn affect their daily life such as eating and sleeping. Scientific and systematic evaluation is the basis and premise for intervention, but there is still a lack of special assessment tools for the oral comfort status of patients with heart failure.

In some studies, a visual analogue scale (VAS numeric score) was used to provide a rough assessment of a patient’s oral comfort ^[7]^, but this method did not provide insight into the specific manifestations and extent of oral discomfort. In addition, some oral assessment tools, such as the Oral Care Assessment Form, indirectly understand the comfort status by assessing the degree of abnormality of the oral anatomy, and do not directly reflect the personal feelings of the patient. Kolcaba’s Theory of Comfort states that comfort is a comprehensive state that involves four dimensions: physiological, psychological, social, and environmental. This theory provides a theoretical basis for a comprehensive assessment of the patient’s comfort status, especially in the assessment of subjective feelings. Based on this theory, the patient’s subjective experience can be used to better understand the manifestation and extent of oral discomfort. Keeping comfort as a goal of care can help to develop individualized interventions that improve patients’ quality of life. Through literature review, expert consultation and questionnaire survey, combined with Kolcaba’s theory, a comprehensive and scientific evaluation framework was constructed. The reliability and validity of the questionnaire were tested to ensure its reliability and validity in practical application. The tool can dynamically and scientifically evaluate the effectiveness of oral care interventions, helping healthcare professionals to develop precise care plans to improve the oral comfort of HF patients.

In this study, a questionnaire for the assessment of oral comfort in hospitalized patients with heart failure was developed through literature review, expert consultation and questionnaire survey, and the reliability and validity test were carried out, in order to provide an effective tool for the oral comfort assessment of HF patients in clinical nursing.

## Material and methods

### Tool development

The questionnaire was developed through four key steps: establishing an item pool, conducting expert consultations, performing a pre-experiment, and carrying out clinical testing.

### Consulting with an expert and survey participants

Criteria for experts consulted via Letter: (1) Bachelor’s degree or above; (2) Nurse in charge or above; (3) Engaged in basic nursing, oral care or cardiovascular nursing for more than 10 years, knowing relevant knowledge of research; (4) Voluntary participation in this study.

#### Inclusion criteria

(1) Hospitalized patients with an age ≥ 18 years and a diagnosis including heart failure; (2) New York cardiac function grade II ∼ IV; (3) Clear consciousness and ability to communicate; (4) Informed consent.

#### Exclusion Criteria

(1) Oral tumors, serious oral diseases and other diseases that lead to bias in the survey results. A total of 190 patients with heart failure in three general hospitals (including 2 tertiary hospitals) from June 2020 to September 2020 were finally enrolled (Table 2).

### Preliminary development of the questionnaire

Based on Kolcaba’s comfort theory ^[8]^, comfort is mainly embodied in four aspects: physical, psychological, social, and environmental, combined with Locker’s oral health measurement theory ^[9-12]^. Based on interviews conducted with inpatients suffering from heart failure, a comprehensive framework for assessing oral comfort was developed, encompassing four key dimensions: physical comfort, psychological comfort, social comfort, and environmental comfort. This framework includes 10 primary areas, which are further broken down into 36 specific sub-items. These areas reflect the multifaceted nature of oral comfort, particularly in the context of heart failure, where both oral health and overall well-being are significantly impacted. Each item is scored on a 5-point scale, with higher scores indicating greater discomfort.

Six specialists in primary care, oral care, or cardiovascular care were selected for consultation. The expert consultation form was divided into two parts, the first part is the expert opinion consultation on the questionnaire dimension and item setting, and the second part is the basic information of the expert. The first part briefly explains the dimensions of the questionnaire and the theoretical basis for the setting of the items, and asks experts to make judgments on the rationality of the listed items and the relevance of the items to the questionnaire based on their personal practical experience, personal judgment or reference to domestic and foreign literature, relevant theoretical basis, etc., and put forward suggestions for revision. Rationality was divided into two judgments: “reasonable” and “unreasonable”, if “reasonable” is selected, the relevance of the items is further judged, and the relevance is divided into “unrelated”, “weakly correlated”, “relevant” and “strongly correlated”, and the corresponding scores are 1, 2, 3 and 4. The second part of the basic information of experts mainly included the basic information of experts, the familiarity of experts, and the basis for expert judgment.

The research group sorted out the relevant materials and send them to the consulted experts by email, and ask the experts to reply within one week, and sort out the data of the round of expert consultation after receiving the reply. After an interval of one week, the second round of consultation questionnaires was sent to the experts, and the second round was same as the first round, and the expert consultation was terminated when the expert coordination coefficient (Kendall’s W) > 0.5. In the first round of expert consultation, the mean correlation value ≥ 3 points, the coefficient of variation ≤0.4, and the expert judgment “reasonable” were used as the screening indicators. In the second round of expert consultation, the mean correlation value ≥ 3 points, the coefficient of variation ≤0.34, and the expert judgment “reasonable” were used as the screening indicators, and the items that met all the above indicators were retained, the items that did not meet two or more indicators were deleted, and the items that did not meet one index were decided to be deleted through group discussion to form a pre-survey version of the questionnaire.

#### Pre-survey

A pre-survey of 20 heart failure patients who were hospitalized in the cardiology department of a tertiary hospital in July 2020 was conducted by suitability sampling, with the purpose of understanding whether the inpatients could understand the content of the questionnaire and answer, with an average filling time of 7 minutes. According to the results of the pre-survey, the expression of some items was modified, and the clinical version of the “Questionnaire on Oral Comfort of Inpatients with Heart Failure” was finally formed.

#### Clinical evaluation of questionnaires

The survey included general data and the “Oral Comfort Assessment Questionnaire for In patients with Heart Failure”, and the modified Beck Oral Assessment Scale (BOAS) was selected as the standard validity control. The sample size of the questionnaire reliability and validity test is generally 5∼10 times the number of items, and there were 32 items in the clinical test version, with a sample size of 160∼320 cases, and a total of 192 questionnaires were issued, and 190 valid questionnaires were recovered, with an effective recovery rate of 98.9%. The critical ratio decision value (CR)^[13]^ was used to screen the items, and the upper and lower 27% of the questionnaire score ranking were used as the cut-off value, and the patients were divided into high and low groups, and the differences between the two groups in each item were compared by independent sample t-test, and the test level was α=0.05, and the items that did not reach the significant level of the t-test were deleted.

### Statistical methods

The data were carefully verified for accuracy and analyzed using SPSS version 18.0 software. During the expert consultation stage, various metrics were examined, including the expert authority coefficient (Cα), questionnaire coordination coefficient (Kendall’s W), Cronbach’s α coefficient, concentration degree (mean Mj), and item coefficient of variation (Vj). The reliability of the questionnaire was assessed using Cronbach’s α coefficient and split-half reliability, ensuring a robust evaluation of its internal consistency and dependability ^[14]^. The validity of the questionnaire was tested by using the validity of the validity scale, the validity of the content and the validity of the structure: (1) the validity of the validity scale was measured by the oral Beck score scale, and the Pearson correlation coefficient and significance test of the two tables were analyzed; (2) Content validity was calculated at the item level content validity (I-CVI) and scale level content validity (S-CVI/UA, S-CVI/Ave); (3) Exploratory factor analysis (EFA) was conducted to evaluate the structural validity of the questionnaire. Principal component analysis (PCA) was employed, followed by maximum orthogonal rotation of variance to optimize factor loadings. Common factors were extracted based on an eigenvalue >1 and a cumulative contribution rate >50%. Items with a factor loading <0.4 were excluded, ensuring a robust and well-structured questionnaire design ^[15]^.

### Quality control

The item pool’s build quality control was ensured throughout the process. Two master’s students independently searched, screened, and evaluated the literature, and screened the items. After completing this step, the findings were summarized. In cases of differing opinions, the decisions were made jointly through group discussion. Expert consultation for quality control was conducted after selecting the experts. The significance, purpose, and objectives of the study were explained to the experts via phone or email. Questionnaires were issued after obtaining their consent. The design of the expert consultation questionnaire was finalized through group discussion and commissioning. The interface was kept simple and easy to understand, facilitating ease of completion and providing sufficient blank space for experts to exchange opinions.

Quality control of clinical reliability and validity tests was conducted as follows. A total of 20 patients were selected for pre-investigation, and the questionnaire design and questioning methods were adjusted accordingly. Before testing the clinical reliability and validity of the questionnaire, the consent of the ward’s head nurse was obtained, along with general information on HF patients in the ward. During the investigation, the investigators briefly explained the purpose and significance of the study to the patients. The questionnaire was distributed after obtaining their consent. For patients with vision problems or limited mobility, the researcher assisted in completing the form to ensure accuracy. The validity and data integrity of the questionnaire were checked on the spot during its collection.

## Results

### Characteristics of consulting specialists and patients

Finally, a total of 6 experts from 4 hospitals in 3 provinces were selected as members of the expert group, including basic nursing (2 people), oral care (1 person) and cardiovascular medicine nursing (3 people). Among them, there were 4 master’s degree and 2 bachelor’s degree students, 4 senior students, 1 deputy senior and 1 intermediate level; Age 36∼55 years old, average 45±6.72 years old, engaged in related clinical work for 13∼31 years, average 21±7.05 years (Table 1). A total of 190 patients with heart failure in three general hospitals (including 2 tertiary hospitals) from June 2020 to September 2020 were finally enrolled (Table 2).

**Table 1.** Basic information on experts.

| Project | Classification Number | Number | Percentage (%) |
| --- | --- | --- | --- |
| Sex | Male | 0 | 0 |
|  | Female | 6 | 100 |
| Age | Over 50 years old | 1 | 16.67 |
|  | 40-50 years old | 4 | 66.67 |
|  | Less than 40 years old | 1 | 16.67 |
| Highest Education | Ph. | 0 | 0 |
|  | Master's Degree | 4 | 66.67 |
|  | Bachelor's Degree | 2 | 33.33 |
| Technical title | Chief Nurse | 4 | 66.67 |
|  | Associate Nurse Practitioner | 1 | 16.67 |
|  | Nurse Practitioner-in-Charge | 1 | 16.67 |
| Years of working experience | More than 25 years | 2 | 33.33 |
|  | 15-25 years | 2 | 33.33 |
|  | 10-15 years | 2 | 33.33 |

**Table 2.** Characteristics of the Sample (n=190)

|  |  |  |
| --- | --- | --- |
| Age in years |  |  |
| Mean (SD) =66±12.35 Range 32-87 |  |  |
| Gender | Male | 111(58.4%) |
|  | Female | 79(41.6%) |
| Area | rural areas | 104 (54.6%) |
|  | townships | 35 (18.5%) |
|  | county seat | 18 (9.3%) |
|  | at or above the city level | 33 (17.6%) |
| Diet Type | General Diet | 186(97.9%) |
|  | Semi-fluid diet | 4(2.1%) |
|  | Fluid diet | 0 |
| Smoking status | Yes | 76(40.0%) |
|  | No | 114(60.0%) |
| Alcohol consumption | Yes | 76(40.0%) |
|  | No | 114(60.0%) |
| Heart Function Classification | Grade 2 | 16(8.4%) |
|  | Grade 3 | 146(76.8%) |
|  | Grade 4 | 28(14.7%) |
| Self-care scores | 0-40 | 7(3.7%) |
|  | 41-60 | 35(18.4%) |
|  | 60-99 | 144(75.8%) |
|  | 100 | 4(2.1%) |
| Combined underlying disease | Yes | 167(87.9%) |
|  | No | 23(12.1%) |
| Sample admission diagnosis | Range | Mean (SD) |
| Beck oral scores | 3-13 | 7.96±2.27 |
| Self-care score | 15-100 | 74.7±16. |
| Left ventricular ejection fraction | 14%-70% | 39±13.6% |
| B-type natriuretic peptide(pg/ml) | 77-35000 | 6971.69±7650.54 |
| Systolic blood pressure (mmHg) | 86-181 | 123±22.82 |
| Diastolic blood pressure(mmHg) | 45-124 | 79±14.33 |

### Expert consultation

According to the results of the first round of expert consultation in Table 3, combined with the screening criteria, “daily leisure activities” were deleted, with two items of “interpersonal relationship with patients”. According to the expert opinion, three items were added: “the degree of influence of the current situation of oral comfort on physical discomfort”, “whether the status of oral comfort causes other physical discomfort”, and “assessment of the current situation of self-confidence brought about by oral comfort”, “Oral language-related language comfort assessment” was divided into two items: “oral cavity-related articulation comfort evaluation” and “oral cavity-related language action comfort evaluation”. Revised “oral speaking-related comfort evaluation” to “degree of language limitations”.

**Table 3.** The advices of Experts.

|  | First round of experts' consultations | Second round of experts' advice |
| --- | --- | --- |
| Personal authority factor | 0.75-0.85 | 0.75-0.9 |
| Group authority coefficient | 0.775 | 0.792 |
| Kendall's W | 0.507 | 0.616 |
| Cronbach's $\alpha$ | 0.973 | 0.892 |
| Mj<3 | 4 entries | 3 entries |
| Vj>0.4 | 3 entries | 11 entries |

**Table 4.**
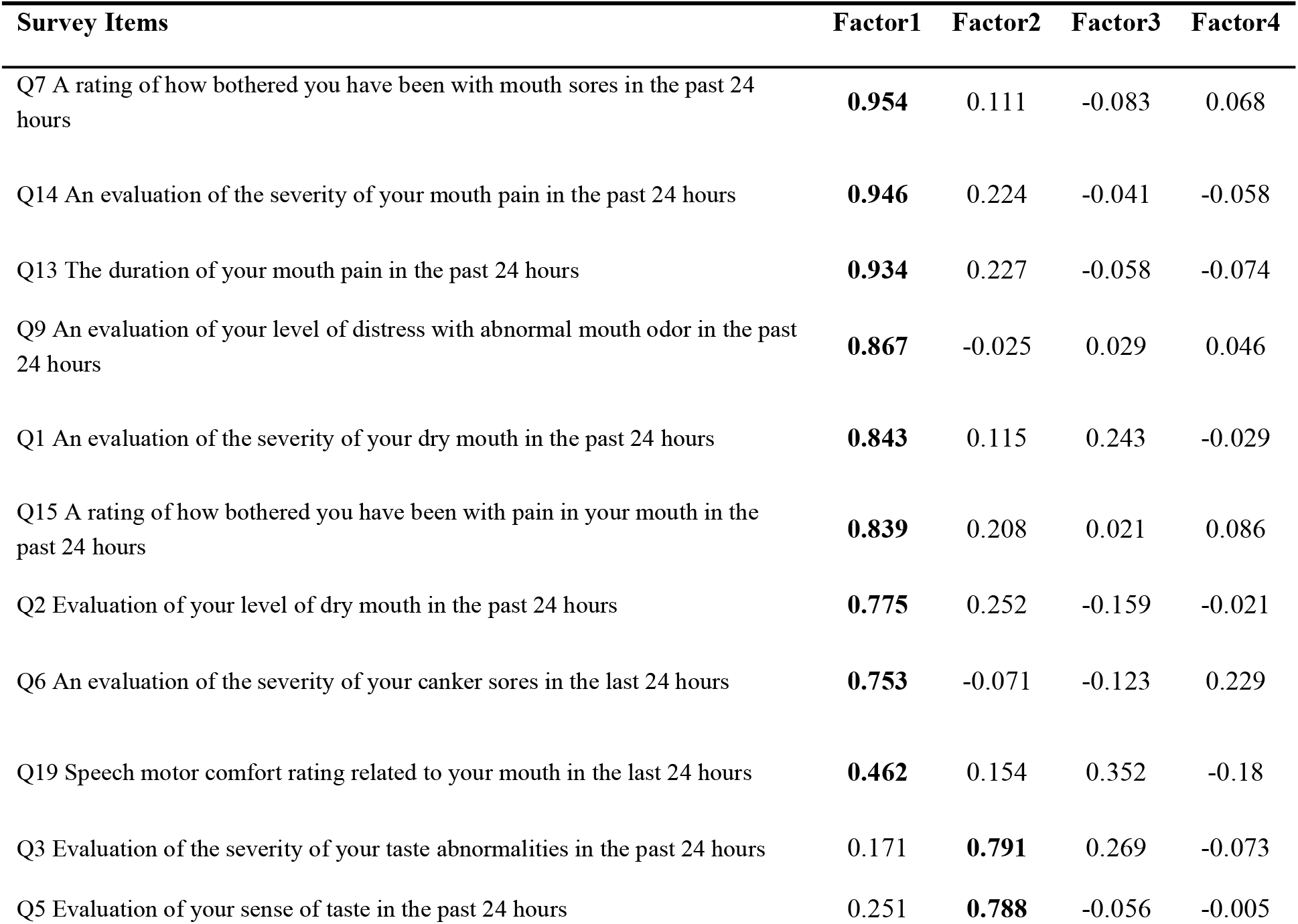
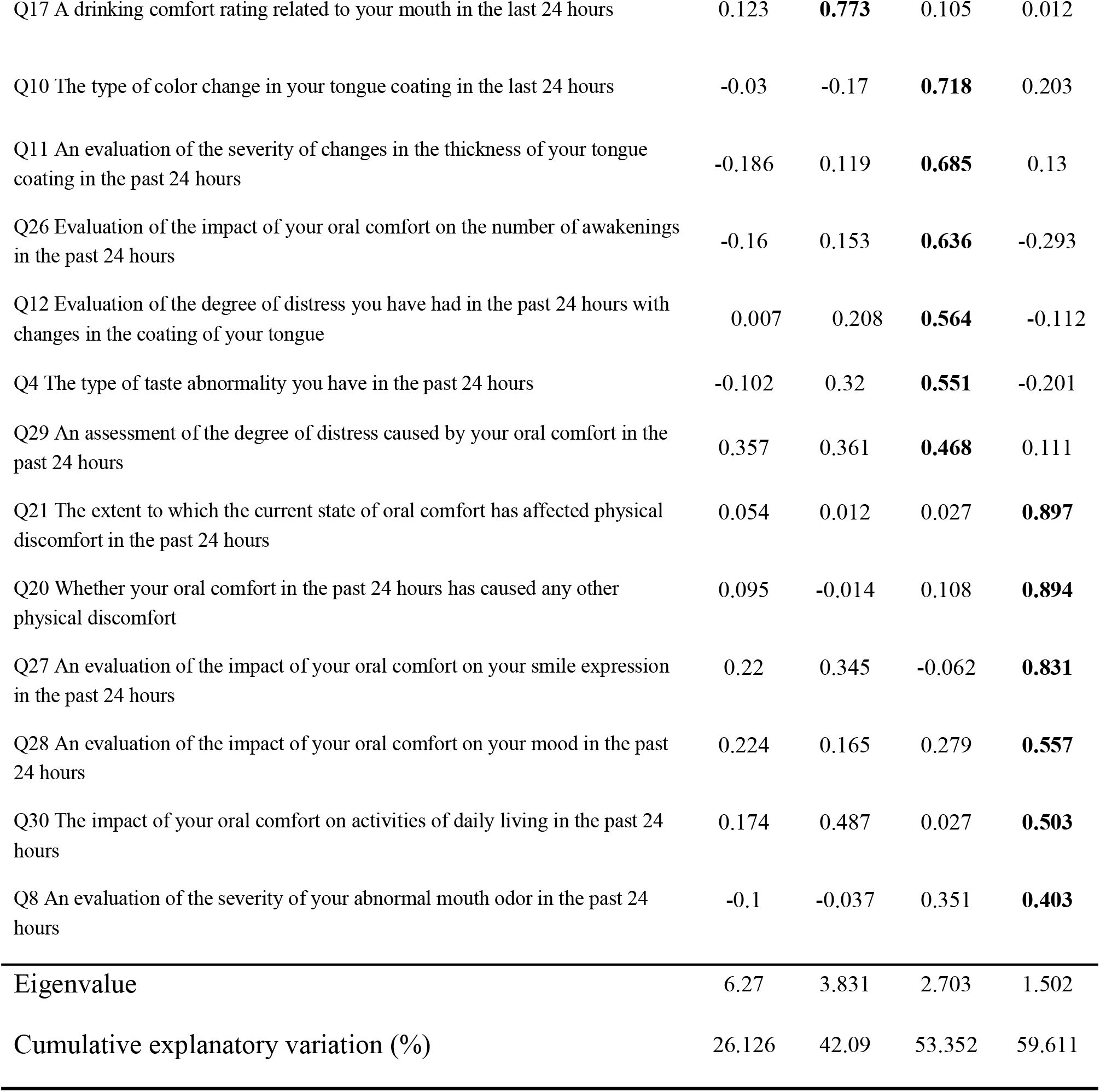
Factor loading matrix.

| Survey Items | Factor1 | Factor2 | Factor3 | Factor4 |
| --- | --- | --- | --- | --- |
| Q7 A rating of how bothered you have been with mouth sores in the past 24 hours | **0.954** | 0.111 | -0.083 | 0.068 |
| Q14 An evaluation of the severity of your mouth pain in the past 24 hours | **0.946** | 0.224 | -0.041 | -0.058 |
| Q13 The duration of your mouth pain in the past 24 hours | **0.934** | 0.227 | -0.058 | -0.074 |
| Q9 An evaluation of your level of distress with abnormal mouth odor in the past 24 hours | **0.867** | -0.025 | 0.029 | 0.046 |
| Q1 An evaluation of the severity of your dry mouth in the past 24 hours | **0.843** | 0.115 | 0.243 | -0.029 |
| Q15 A rating of how bothered you have been with pain in your mouth in the past 24 hours | **0.839** | 0.208 | 0.021 | 0.086 |
| Q2 Evaluation of your level of dry mouth in the past 24 hours | **0.775** | 0.252 | -0.159 | -0.021 |
| Q6 An evaluation of the severity of your canker sores in the last 24 hours | **0.753** | -0.071 | -0.123 | 0.229 |
| Q19 Speech motor comfort rating related to your mouth in the last 24 hours | **0.462** | 0.154 | 0.352 | -0.18 |
| Q3 Evaluation of the severity of your taste abnormalities in the past 24 hours | 0.171 | **0.791** | 0.269 | -0.073 |
| Q5 Evaluation of your sense of taste in the past 24 hours | 0.251 | **0.788** | -0.056 | -0.005 |
| Q17 A drinking comfort rating related to your mouth in the last 24 hours | 0.123 | **0.773** | 0.105 | 0.012 |
| Q10 The type of color change in your tongue coating in the last 24 hours | -0.03 | -0.17 | **0.718** | 0.203 |
| Q11 An evaluation of the severity of changes in the thickness of your tongue coating in the past 24 hours | -0.186 | 0.119 | **0.685** | 0.13 |
| Q26 Evaluation of the impact of your oral comfort on the number of awakenings in the past 24 hours | -0.16 | 0.153 | **0.636** | -0.293 |
| Q12 Evaluation of the degree of distress you have had in the past 24 hours with changes in the coating of your tongue | 0.007 | 0.208 | **0.564** | -0.112 |
| Q4 The type of taste abnormality you have in the past 24 hours | -0.102 | 0.32 | **0.551** | -0.201 |
| Q29 An assessment of the degree of distress caused by your oral comfort in the past 24 hours | 0.357 | 0.361 | **0.468** | 0.111 |
| Q21 The extent to which the current state of oral comfort has affected physical discomfort in the past 24 hours | 0.054 | 0.012 | 0.027 | **0.897** |
| Q20 Whether your oral comfort in the past 24 hours has caused any other physical discomfort | 0.095 | -0.014 | 0.108 | **0.894** |
| Q27 An evaluation of the impact of your oral comfort on your smile expression in the past 24 hours | 0.22 | 0.345 | -0.062 | **0.831** |
| Q28 An evaluation of the impact of your oral comfort on your mood in the past 24 hours | 0.224 | 0.165 | 0.279 | **0.557** |
| Q30 The impact of your oral comfort on activities of daily living in the past 24 hours | 0.174 | 0.487 | 0.027 | **0.503** |
| Q8 An evaluation of the severity of your abnormal mouth odor in the past 24 hours | -0.1 | -0.037 | 0.351 | **0.403** |
| Eigenvalue | 6.27 | 3.831 | 2.703 | 1.502 |
| Cumulative explanatory variation (%) | 26.126 | 42.09 | 53.352 | 59.611 |

According to the results of the second round of expert consultation in Table 2, combined with the screening criteria and few entries were deleted as well, “the severity of your taste abnormalities in the past 24 hours”, “the impact of your oral comfort status on the proportion of awakening time in the past 24 hours”, “the impact of your oral comfort status on your overall sleep quality in the past 24 hours”, “the impact of your oral comfort status on your emotional symptoms in the past 24 hours”, “the impact of your oral comfort status on treatment coordination activities in the past 24 hours”, and “ The impact of your oral comfort in the past 24 hours on your patient’s relationship with your caregiver”, “The impact of your oral comfort in the past 24 hours on your patient’s relationship with your family”. After two rounds of expert consultation, the clinical version of the “Oral Comfort Assessment Questionnaire for Inpatients with Heart Failure” composed of 32 items was finally formed.

### Validity test

#### Validity criteria

In this study, the Beck oral scoring method (BOAS table) was selected as the standard validity control. The results showed that the correlation coefficient between the questionnaire and the BOAS table Pearson was 0.233, P<0.05.

### Content validity

The content validity analysis of the questionnaire includes two key measures: item-level content validity (I-CVI) and scale-level content validity (S-CVI/UA, S-CVI/Ave). The I-CVI of each item of the questionnaire was between 0.833∼1, the S-CVI/UA was 0.875, and the S-CVI/Ave was 0.979, and it was generally believed that the I-CVI must be above 0.78, the S-CVI/UA should not be less than 0.8 points, and the S-CVI/Ave should be above 0.90, indicating that the scale has good content validity.

### Structural Validity

The results revealed that the Kaiser-Meyer-Olkin (KMO) value of the questionnaire was 0.606, and the Bartlett’s sphericity test yielded a Chi-square value of 1895.778 (P < 0.001), confirming that the data were suitable for factor analysis. Principal component analysis (PCA) was used to extract the common factors, with an eigenvalue ≥ 1, and the component matrix was derived using orthogonal rotation with maximum variance. The initial round of factor analysis identified eight common factors with a cumulative variance contribution rate of 74.586%. Items with a factor loading < 0.4 were removed, including:

“Q16: Eating comfort evaluation related to oral cavity in the past 24 hours”

“Q18: Pronunciation comfort evaluation related to oral cavity in the past 24 hours”

“Q24: Evaluation of the influence of oral comfort status in the past 24 hours on the time it takes to fall asleep.”

After further analysis and factor extraction, the rotated factor loading matrix showed that the cumulative variance contribution rate of the first four common factors reached 59.611%, with each item having a factor loading > 0.4. Additionally, the scree plot (Fig. 1) indicated a stable slope after the fourth factor, leading to the final extraction of four common factors. The resulting Oral Comfort Assessment Questionnaire for Inpatients with Heart Failure comprised 4 dimensions and 24 items. The factor loading matrix and questionnaire items are detailed in Table 3, demonstrating strong structural validity.

**Figure 1.**
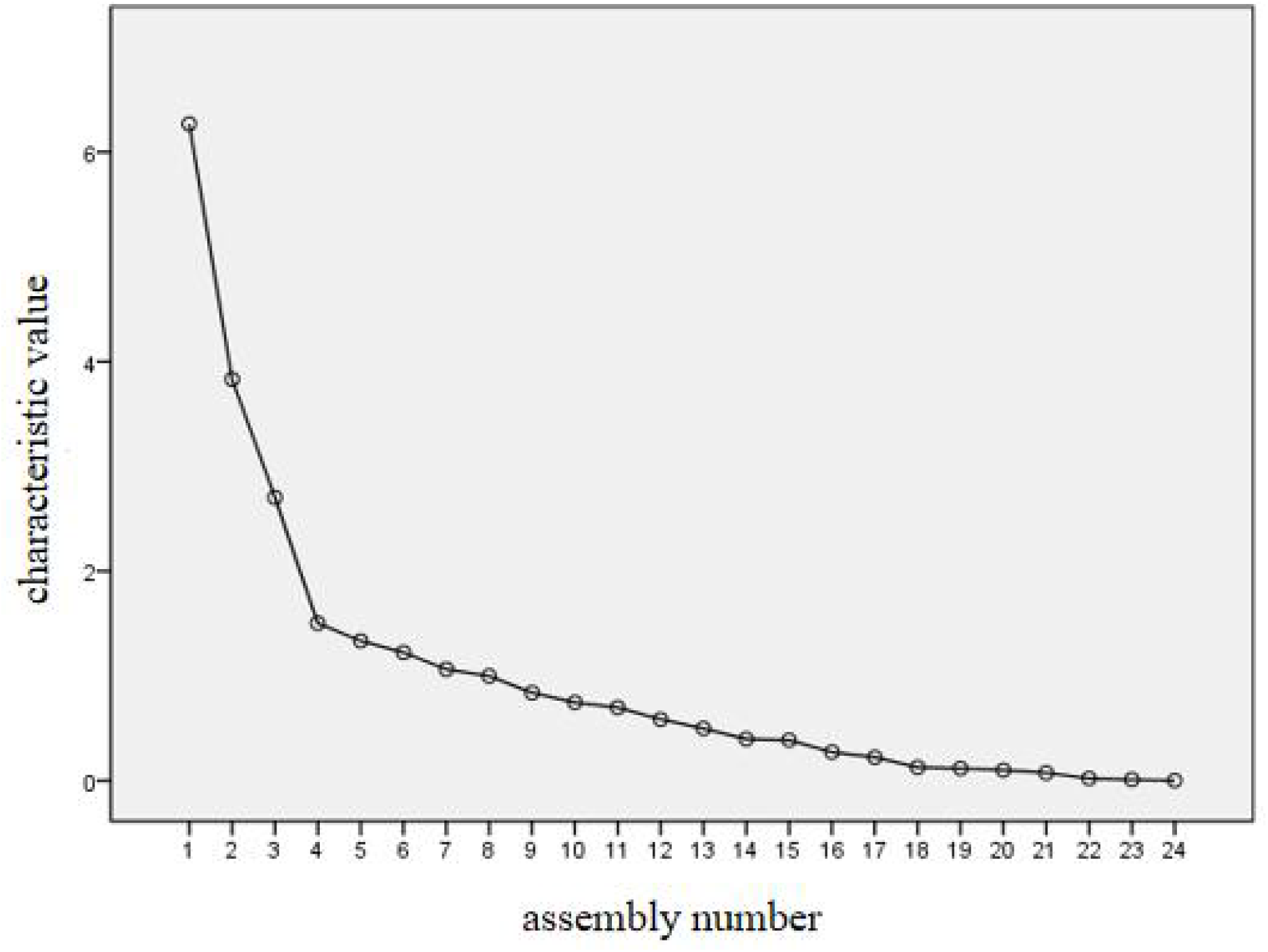
Gravel diagram

### Reliability test

Reliability mainly evaluates the stability or accuracy of the measurement tool and indicates the reliability of the data. The questionnaire used half-folded reliability (Spearman-Brown coefficient) and Cronbach’s α coefficient as reliability test indicators. After statistical analysis, the Cronbach’s α coefficient and the Spearman-Brown coefficient were 0.810 and 0.732, indicating that the questionnaire had good internal consistency.

## Discussion

The questionnaire was developed based on Kolcaba’s Comfort Theory as the guiding theoretical framework, supplemented by relevant literature on heart failure symptom management. Additionally, a range of validated assessment tools was consulted, including the Fridman Heart Failure Symptom Checklist, the Cardiac Symptom Survey (CSS), the Anderson Symptom Inventory – Heart Failure, and the Memorial Symptom Assessment Scale for Heart Failure, among others. These resources were used to construct a comprehensive pool of questionnaire items that address various symptoms and aspects of comfort in heart failure patients. During the compilation of the questionnaire, relevant nursing experts from 3 provinces and 4 hospitals conducted rigorous screening and judgment of the questionnaire items through two rounds of Delphi expert consultation, and put forward detailed professional opinions on the content of the questionnaire. Through clinical testing, the questionnaire had good reliability and validity, and can be applied to the oral comfort assessment of HF patients, so as to provide a valuable reference for clinical nursing practice.

A well-structured questionnaire that elicits good patient responses is essential for effective data collection. Reliability refers to the stability and consistency of a measurement instrument over time. One common method for evaluating reliability is split-half reliability, which involves dividing the survey items into two equal parts and calculating the correlation coefficient between the scores of these two halves. This approach is considered an internal consistency coefficient and is widely used in the reliability analysis of attitude and opinion questionnaires. By assessing the correlation between the two halves, this method provides insight into how consistently the instrument measures the intended construct. The Spearman-Brown coefficient is a widely used statistic in this type of analysis. For the questionnaire in question, the Spearman-Brown coefficient was found to be 0.732, which falls within the acceptable range for reliability, indicating that the instrument exhibits reasonable internal consistency.

Additionally, the Cronbach’s α (alpha) reliability coefficient was used to assess the overall internal consistency of the questionnaire. Cronbach’s α represents the average of the half-split reliability coefficients of all the items in the questionnaire and is one of the most commonly used methods for evaluating reliability. Generally, a Cronbach’s α coefficient within the range of 0.7 to 0.8 is considered acceptable. A value in the range of 0.8 to 0.9 suggests good internal consistency, while a value greater than 0.9 indicates very good consistency. In this case, the Cronbach’s α coefficient for the questionnaire was found to be 0.810, which indicates that the questionnaire has good internal consistency. This suggests that the items in the questionnaire are reliably measuring the same underlying construct, making it a reliable tool for assessment.

Validity is the basis for whether the response measurement tool is scientific, mainly including standard validity, content validity, and structural validity. Criterion validity is to select a tool as the calibration criterion and analyze the relationship between the questionnaire and the calibration criterion. In this study, the Beck Oral Score was selected to evaluate the basic physiological status of the oral cavity, which is an objective assessment method, and the nursing staff mainly evaluated and scored the oral physiological status of the patients. The Pearson correlation coefficient between the Oral Status Assessment Questionnaire for Hospitalized Patients with Heart Failure and the score was 0.233 and P<0.05, indicating that there was a weak correlation between the two. The reason for the analysis is that the Beck oral score is an objective scoring method, and the “Oral Status Assessment Questionnaire for Inpatients with Heart Failure” is a subjective scoring method, which not only includes the assessment of the discomfort caused by the patient’s oral physiology, but also includes the psychological and social mapping of oral discomfort to oneself. Content validity refers to the ability of the items in the measurement tool to reflect the measurement content, which is generally explained by the method of expert evaluation, and this study refers to the recommendation of “Statistical Methods and Software Operation Practice of Nursing Research” ^[15]^, which selected 6 relevant experts for evaluation. The I-CVI of each item of the questionnaire was 0.833∼1, and the S-CVI/UA was 0.875 and S-CVI/Ave was 0.979. It is generally believed that the I-CVI must be above 0.78, the S-CVI/UA score should not be less than 0.8, and the S-CVI/Ave should be above 0.90, indicating that the scale had good content validity. The cumulative variance contribution rate of exploratory factor analysis of the questionnaire was 59.611%, and the factor load of each item was > 0.4, indicating that the structure validity of the debugged questionnaire was reliable.

## Conclusion

The Oral Comfort Assessment Questionnaire for Inpatients with Heart Failure developed in this study is grounded in Kolcaba’s Comfort Theory and demonstrates strong reliability and validity. This tool is effective for assessing the oral comfort of hospitalized heart failure patients in China, providing a quantitative basis for evaluating their oral comfort. It also serves as an essential resource for nursing staff to gain a deeper understanding of the oral comfort needs of these patients.

Given that this is a self-assessment questionnaire reflecting patients’ subjective experiences, it is recommended to be used alongside objective clinical indicators of oral physiology. This combination allows for a more comprehensive evaluation of oral comfort. By integrating both subjective and objective assessments, healthcare providers can gain a thorough understanding of the oral comfort status of heart failure patients, facilitating improved care and more targeted interventions.

## Limitations

In this study, convenience sampling was used to recruit research subjects, and the representativeness of research subjects was limited. Due to the cost expenditure, some research indicators were only selected before the intervention and on the 7th day of the intervention for comparison, and were not included. Continuous observation does not allow for in-depth analysis of the changes in the research indicators. Due to time constraints, this study only carried out a 7-day intervention during hospitalization for clinical patients, and none method to explore the medium- and long-term effects of the intervention.

## Data Availability

All relevant data are within the manuscript and its Supporting Information files.

## Author contributions

**Conceptualization:** Mei Lu, Runyu Yang , Ming Tao.

**Methodology:**Mei Lu, Runyu Yang, Xiaona Li,Xinkun Wang, Ming Tao.

**Project administration:** Ming Tao.

**Supervision:** Mei Lu.

**Writing – original draft:** Runyu Yang.

**Writing – review** & **editing:** Mei Lu, Runyu Yang , Ming Tao.

## Author Information

**First author:** Mei Lu, Cardiovascular Nurse, B.S

**Affiliation:** Department of Cardiovascular Medicine, Affiliated Hospital of Zunyi Medical University, School of Nursing, Zunyi, Guizhou, Zunyi, Guizhou, China.

**Address:** No. 149, Dalian Road, Huichuan District, Zunyi City, Guizhou, China, Postal Code: 563000

**Co-first authors:** Runyu Yang, RN, Master

**Affiliation:** Department of Hematology, West China Hospital, Sichuan University, Chengdu, Sichuan, China/ West China School of Nursing, Sichuan University, Chengdu, Sichuan, China.

**Address:** PO Box 610041, No.37 Guo Xue Street, Chengdu, Sichuan Province, P.R. China

**Author:** Xiaona Li, master’s degree candidate

**Affiliation:** School of Nursing, Zunyi Medical University, Guizhou, China.

**Address:** No. 149, Dalian Road, Huichuan District, Zunyi City, Guizhou, China **Email:**

**Author:** Xinkun Wang, master’s degree candidate

**Affiliation:** School of Nursing, Zunyi Medical University, Guizhou, China.

**Address:** No. 149, Dalian Road, Huichuan District, Zunyi City, Guizhou, China

**Corresponding Author:** Ming Tao, Director of Nursing Department, Graduate Supervisor, Professor

**Unit:** Nursing Department, Affiliated Hospital of Zunyi Medical University, Guizhou

**Address:** No. 149, Dalian Road, Huichuan District, Zunyi City, Guizhou, China

We would like to thank Prof. Ling Chen, Prof. Xueqin Gao, Prof. Xinglian Shi, Prof. Wenbi Yang, and Prof. Changxiu Li for their analysis and review of the scale. We are also grateful to all the patients and researchers who participated in this study.

## Data Availability Statement

Data supporting the results of this study may be made available upon reasonable request by the corresponding authors.

## Ethical considerations

This study was reviewed and approved by the Biomedical Research Ethics Committee of the Affiliated Hospital of Zunyi Medical College, approval number: KLLY-2019-040. For study purposes, we obtained written informed consent from all patients.

## Disclaimer

The views expressed in the submitted articles are the author’s own and not the official position of the institution and funders.

